# The Impact of Pregnant Women’s Dietary Behavior on the Physiological Adaptation Paradox and Maternal-Fetal Resource Conflict in Conflict Settings: A Predictive Analytical Study

**DOI:** 10.64898/2026.06.17.26355845

**Authors:** Muneer Musleh Al-Wesabi, Nageeb Ali Eskander

**Affiliations:** Al-Razi University, 21 September University for Medical and Applied Sciences, Sana’a, Yemen; Al-Razi University, Sana’a, Yemen

**Keywords:** nutritional awareness, dietary practices, health indicators, antenatal care, fetal programming, Sana’a Capital Governorate, Yemen

## Abstract

This scientific study aims to assess the level of awareness, nutritional knowledge, and actual behavioral practices among pregnant women in the Capital District of Sana’a, Republic of Yemen, and to determine their impact on the health and clinical indicators of the mother and fetus under complex conflict conditions. The study employed a descriptive-analytical approach based on a simple random sample of 200 pregnant women attending government-run hospitals and specialized medical centers in the Capital District. Field data were collected during December 2025 using a structured and validated questionnaire consisting of 42 items measuring demographic variables, awareness, practices, barriers, and health outcomes.

The results of the statistical analysis using SPSS software showed a high level of nutritional awareness (87%) and healthy dietary practices (80%) among the sample participants. Simple and multiple linear regression tests revealed a statistically significant effect of awareness and practices in explaining 20.2% of the variance in the health status of the mother and fetus (^R²^= 0.204, *p* < 0.001). The study demonstrated that actual behavioral practices have greater predictive power (β=0.316, p=0.001) compared to theoretical cognitive awareness (β=0.232, p=0.005) in determining clinical outcomes for the mother and fetus, highlighting the widening gap between knowledge and behavior under structural pressures. “Morning sickness” (80%) and the deterioration of “family economic status” (71%) emerged as the greatest physiological and material barriers to proper nutrition. With their inferential impact established as an extension of the maternal-fetal resource allocation conflict in a physiologically and economically challenging environment, the study also identified significant differences in nutritional behavior and health outcomes in favor of housewives and mothers who are more educated and have higher incomes, while no significant differences were recorded attributable to obstetric variables such as stage or order of pregnancy.

The study offers a unique theoretical and practical contribution by formulating an integrated causal model that demonstrates that the fetus acts as a biological drain on the mother’s cellular and mineral reserves in a war environment, which necessitates directing antenatal care and support programs toward effective behavioral empowerment and nutritional support to overcome the structural and material barriers faced by pregnant women.

**Highlights:**

- The nutritional awareness and practices of pregnant women account for 20.4% of the total variance in the combined health status of the mother and fetus in a conflict environment.
- Applied behavioral practices have a stronger predictive effect (β=0.316) on health indicators compared to theoretical cognitive awareness (β=0.232).
- Objective fetal indicators show relative stability at the expense of a sharp decline in the mother’s subjective and physical indicators due to the mechanism of physiological adaptation within the uterus.
- Economic determinants and low purchasing power dominate as the strongest structural barriers to dietary diversity, with significant statistical differences indicating that these obstacles are compounded among low-income pregnant women.
- The activation of the maternal-fetal resource allocation conflict as a forced physiological adaptation mechanism to ensure species survival in conflict environments.
- The fetus acts as a “distinct biological drain” that draws on the mother’s stored cellular hydrocarbons and minerals, which explains the deterioration of her physical and subjective psychological indicators in contrast to the stability of her fetus’s apparent growth.
- Previous obstetric experiences (pregnancy order and stage) have no statistical effect on the mother’s awareness and practices, necessitating the provision of standardized educational programs for all pregnant women.

## 1. Introduction

Proper nutrition for the pregnant mother during pregnancy is considered the most significant biological and physiological determinant of the health status of successive generations, as this brief period constitutes the pivotal link in determining the newborn’s quality of life and future physical and mental health (Abu-Saad & Fraser, 2010). Healthy nutrition for pregnant women is defined as a balanced and comprehensive intake of macronutrients such as carbohydrates, proteins, and fats, and micronutrients such as vitamins and minerals—including folic acid, iron, and calcium—with the aim of supporting cell division, placental development, and the formation of the fetus’s vital organs without depleting the mother’s biological and cellular reserves (Mousa et al., 2019). Recent medical studies indicate that nutritional imbalances—whether excess or deficiency—lead to disruptions in fetal programming for disease, exposing the fetus in the long term to risks of chronic diseases in adulthood, a physiological concept rooted in Barker’s hypothesis on the developmental origins of health and disease (Barker, 1995; Horne et al., 2024).

At the international and regional levels, malnutrition and anemia pose significant challenges to reproductive public health, with nearly 40% of pregnant women worldwide suffering from iron-deficiency anemia, and this rate rises to over 60% in developing and conflict-affected countries (Global Nutrition Report, 2022). In this complex context, the mother’s nutritional awareness emerges as a critical factor in prevention; indeed, the mother’s possession of the information and skills necessary to understand medical guidelines and select locally available foods forms the foundation for shaping her daily dietary behavior (Oladebo et al., 2025; Vidgen & Gallegos, 2014), However, the knowledge-behavior relationship remains subject to numerous structural, economic, and logistical pressures that may prevent the translation of theoretical knowledge into actual practice, resulting in what is known in health psychology as the “knowledge-behavior gap” (Rajaeieh et al., 2026).

In contemporary Yemen, this issue has taken on extremely serious and tragic dimensions as a result of the conflict that has been ongoing for more than a decade, which has led to the disruption of nearly 40% of medical facilities and caused a near-total collapse of infrastructure and household purchasing power (Al-Wesabi, 2020; Healthy Newborn Network, 2024). The 2026 Humanitarian Needs Assessment reports that Yemen faces one of the world’s worst crises of food insecurity and acute malnutrition, with nearly half of children under five suffering from stunting and more than 1.3 million pregnant and lactating women expected to require urgent therapeutic interventions for severe malnutrition (Perez Duque et al., 2025; UNICEF, 2025; UNFPA, 2025). As a densely populated urban center, the capital city of Sana’a reflects the interplay of severe economic poverty with misguided nutritional beliefs and negative traditional practices in shaping the difficult clinical reality for mothers and their fetuses (Al Mansoob & Masood, 2019; Haza’a, 2022).

### Research Problem

The research problem of the current study lies in the continuous deterioration of nutritional health indicators among pregnant women in the Capital District of Sana’a, where food scarcity and availability are no longer the sole obstacle, but is compounded by a widening gap between awareness and practice, as well as the prevalence of harmful traditional beliefs in an environment lacking integrated institutional nutrition education (Al-Wesabi, 2012; Demurtas & Occansey Agbeko, 2025). The Global Nutrition Report notes that Yemen has completely deviated from the international trajectory for achieving the goals of reducing anemia among women of reproductive age, which affects 61.5% of them. This places a heavy biological and clinical burden on nursing and midwifery practices through a doubling of cases of severe postpartum hemorrhage, preeclampsia, and the birth of preterm and low-birth-weight infants (Asferie et al., 2025; Global Nutrition Report, 2022; UNICEF, 2025).

Conceptually, the problem lies in the inadequacy of traditional guidance models that focus on providing medical information to mothers without addressing the structural, material, and physiological barriers they face daily amid conditions of war and poverty (Abas et al., 2020). Although there are Yemeni studies that have addressed the prevalence of anemia or general knowledge about the risks of toxoplasmosis infection (Abdul-Ghani et al., 2024), there is a clear scarcity and knowledge gap in quantitative, inferential, and analytical studies that predict the impact of awareness and practices as independent variables on perceived maternal and fetal health indicators as dependent variables, while measuring the scale and obstacles of reality in the contemporary Sana’a environment of 2026.

### Research Questions

The study seeks to answer the following main question: **What is the impact of the level of awareness and dietary practices of pregnant women in the Capital District of Sana’a on perceived health indicators for the mother and fetus?** The following sub-questions stem from this main question:

- What is the level of nutritional awareness and culture, and the extent to which prevailing behavioral practices are adopted by pregnant women in the Capital District of Sana’a?
- What is the magnitude and direction of the predictive statistical effect of nutritional awareness and practices (collectively and individually) on perceived health indicators for the mother and fetus?
- What is the nature and extent of the economic, physiological, and social barriers faced by pregnant women, and how do they vary according to the mother’s socio-economic determinants?
- Do levels of awareness, practices, and perceived health indicators for the mother and fetus vary according to the mother’s demographic, functional, and obstetric variables?

### Study Objectives

The study aims primarily to achieve the following:

- To measure the overall predictive impact of dimensions of nutritional awareness and practices on perceived health indicators for the mother and fetus in conflict settings.
- To assess the nutritional awareness of pregnant women in Sana’a regarding food groups, micronutrients, and the risks of harmful local dietary behaviors.
- To monitor actual dietary practices (dietary diversity, food hygiene, and adherence to medical supplements) and assess their alignment with established health standards.
- Identify the nature and extent of material, physical, and social barriers faced by pregnant women and analyze their differences and distribution according to their economic and demographic characteristics.
- Examine and identify statistically significant differences in mothers’ responses based on their demographic, occupational, and obstetric characteristics to provide accurate policy recommendations.

### Scientific and Practical Significance of the Research

**· Scientific (Theoretical) Significance:** This lies in providing a sound and innovative academic reference for nursing and midwifery sciences that predicts the impact of cognitive and behavioral variables on perceived health indicators for the mother and fetus, while testing the suitability and validity of health psychology models (such as social cognitive theory and the health belief model) in explaining the behaviors of pregnant women under the severe pressures of economic crises and prolonged wars.

**· Practical (Applied) Significance:** This involves providing the Ministry of Public Health and Population, and international humanitarian organizations (such as UNICEF, the World Health Organization, and the United Nations Population Fund) with up-to-date field and statistical data () for the year 2026, which will help direct aid, supplementary packages, and educational interventions toward the most vulnerable and materially and cognitively deprived groups and mothers in Sana’a.

## 2. Research Hypotheses

Based on the research questions and the problem statement, the following statistical hypotheses were formulated and tested:

• **Main Hypothesis 1 (H1):** There is a statistically significant effect at the significance level (α ≤ 0.05) of pregnant mothers’ nutritional awareness and practices on improving perceived health indicators for mothers and fetuses in the Capital Secretariat of Sana’a. Derived from this are:

**`o` First Sub-Hypothesis (H1.1):** There is a statistically significant effect of nutritional awareness and culture on improving perceived health indicators for the mother and fetus.

**`o` Second sub-hypothesis (H1.2):** There is a statistically significant effect of dietary practices and behaviors on improving perceived health indicators for the mother and fetus.

**`o` Sub-hypothesis 3 (H1.3):** The predictive power and relative contribution of the independent variable’s dimensions differ when they are entered together into a multiple regression model, with actual behavioral practices having greater predictive weight (higher β) in determining clinical outcomes compared to theoretical cognitive awareness.

• **Second Main Hypothesis (H2):** There are statistically significant differences at the significance level (α ≤ 0.05) in the levels of awareness and the extent of encountering barriers (economic, physiological, and social) attributable to the socio-economic determinants of the pregnant mother, particularly the family’s monthly income category, favoring the most disadvantaged groups.

• **Third main hypothesis (H3):** Statistically significant differences at the significance level (α ≤ 0.05) were found in the levels of awareness, practices, and health indicators of the mother and fetus attributable to the mother’s demographic and occupational variables (age, education, monthly income, employment status), while there are no statistically significant differences attributable to obstetric variables (current stage of pregnancy and birth order).

## 3. Theoretical Framework & Literature Review

### Philosophical and Theoretical Foundations of the Cognitive Model

The philosophical and biological grounding of this study stems from the premise that a mother’s dietary behavior during pregnancy is not an isolated, discretionary choice, but rather the product of a continuous, bidirectional interaction between individual cognitive awareness, surrounding material and structural conditions, and the shared physiological clinical outcomes of the mother and her fetus (Mohammad et al., 2025; Rajaeieh et al., 2026). To explain this complex behavioral interaction, the present study draws on three leading explanatory models in health and social psychology:

First, Professor Albert Bandura’s **Social Cognitive Theory (SCT)**, which posits that a pregnant woman’s nutritional behavior is determined through a mechanism of “reciprocal determinism” among three key determinants: the mother’s cognitive and personal factors (such as her awareness, nutritional literacy, and confidence in her ability to change), environmental factors surrounding her (such as food availability, financial capacity, and family and social support), and actual behavior (daily dietary patterns and choices) (Torkan et al., 2018). The theory emphasizes that merely providing theoretical information to the mother will not result in sustainable healthy behavior unless it is accompanied by fostering the mother’s “self-efficacy” and helping her overcome material barriers, as well as providing channels for direct social support within her immediate living environment (SimplyPsychology, 2023).

Second, **the Health Belief Model (HBM)**, which focuses on shaping the pregnant woman’s preventive behavior based on her self-perception of four key physiological and behavioral dimensions (Riazi et al., 2024): perception of risk and threat (such as her awareness of the severity of the risk of the fetus developing neurological defects or stunting due to malnutrition), perception of the severity and difficulty of the consequences (such as the consequences of anemia and severe postpartum hemorrhage on her physiological well-being), and perception of expected preventive benefits (such as adherence to iron supplements and calcium sources to protect herself and her newborn), and comparing them with the perception of existing barriers and disincentives to the behavior (such as financial hardship, symptoms of nausea and morning sickness, and pressures from negative social traditions). The model illustrates that the prevalence of “perceived barriers” and their predominance over “ ” directly leads to the disruption of preventive and healthy nutritional behaviors, which explains the persistence of the behavioral gap among poor pregnant women despite their possession of good levels of cognitive awareness (Rajaeieh et al., 2026).

Third, **the Empowerment Model in Midwifery**, which emphasizes that effective maternal and child care requires transforming the medical relationship from a dry, directive one based on the midwife issuing orders and medical advice, to a genuine partnership aimed at empowering the pregnant woman and equipping her with practical skills, tools, and affordable, locally available nutritional alternatives, thereby strengthening her capacity to make sound health decisions and overcome physical, logistical, and physiological obstacles throughout the journey of pregnancy, childbirth, and beyond.

### A systematic review of previous studies

#### Local (Yemeni) studies

- **Study** (Alsaeedi & Al-Haddad, 2026): This study examined factors influencing the utilization of antenatal care (ANC) services at primary care centers in Sana’a among a representative sample of 422 women. It found that service utilization declined sharply in rural areas (7.6%), It also demonstrated that physical and financial barriers were the major obstacle (55%), and that only 46% of mothers received educational information on proper healthy dietary practices, revealing a severe deficiency in nutritional counseling at current prenatal clinics.
- **Study (**Abdul-Ghani et al., 2024): This study aimed to assess knowledge, attitudes, and practices (KAPs) regarding toxoplasmosis among 410 pregnant women attending antenatal clinics in Sana’a. The results revealed that only 53.3% of the pregnant women were aware of the sources and causes of transmission of the infection, which directly threatens the fetus and causes neurological abnormalities and intrauterine death. The study identified a significant gap in awareness regarding transmission mechanisms via raw meat or unpasteurized dairy products, recommending that food safety be routinely integrated into midwifery care.
- **Study (**Haza’a, 2022): This study examined the knowledge, attitudes, and practices of pregnant women regarding antenatal care in government hospitals in Sana’a (Al-Sabeen and Al-Thawra) among a sample of 371 women. It concluded that 79% had good general awareness, however, actual practices were moderate among 42% of the sample, with a strong positive correlation between educational level and the quality of dietary behavior. The study called for enhancing midwives’ communication and behavioral persuasion skills to provide effective and persuasive nutritional counseling to mothers.
- **Study (**Alsebaeai et al., 2025): This study investigated the prevalence of Pica disorder and its risk factors among 195 pregnant women in Sana’a, finding that the prevalence of this disorder was 28.7%; mothers consumed non-food items such as ice (12.8%), clay (4.6%), petroleum, and ash. Statistical analysis confirmed a close and direct association between Pica and the presence of anemia, iron and zinc deficiency, and increased levels of anxiety and psychological stress associated with prolonged conflict, warning of the severe risks of Pica on nutrient absorption and healthy fetal development.

### Arab, Regional, and International Studies

- **Study (**Rizk et al., 2024): Assessed nutritional knowledge and practices among pregnant women—both local residents and refugees—in Lebanon, and demonstrated that awareness of important micronutrients such as omega-3, iodine, and iron is at its lowest levels (28.4%), with only (20.8%) of participants able to correctly identify iron-rich foods, highlighting the importance of designing educational programs aimed at clarifying the nutritional content of locally available and affordable foods.
- **Study** (Rajaeieh et al., 2026): Conducted the first comprehensive assessment of nutritional knowledge, attitudes, and practices among pregnant women using a large sample of 1,535 participants. The results showed that college-educated mothers had 3.2 times better awareness and practices compared to those without formal education. It also identified deteriorating economic conditions as a critical factor and barrier preventing families from achieving proper dietary diversity and securing balanced meals.
- **Study (**Riazi et al., 2024): This study evaluated the impact of a nutrition education program based on the Health Belief Model (HBM) on dietary intake among pregnant women using a quasi-experimental design. and the results demonstrated that targeting the pregnant woman’s individual perception of the severity of anemia and the benefits of preventive behavior had a direct and statistically significant effect on the quantity and quality of food actually consumed and adherence to medical supplements, thereby supporting the adoption of theoretical behavioral models in healthcare programs to ensure the sustainability of behavioral change among mothers.

### Comprehensive Critical Review of Previous Studies

Through a careful and in-depth review of the previous literature, the researchers note that most medical and clinical studies focus on monitoring the prevalence rates of anemia and fetal malformations as physiological conditions isolated from the pregnant mother’s complex behavioral and cognitive systems, while behavioral studies have been limited to measuring knowledge, attitude, and practice (KAP) indicators in a dry, descriptive manner that does not link these behavioral dimensions to the immediate clinical and biological outcomes and indicators for the mother and her fetus within a unified causal model (Al-Rabeei et al., 2023; Paraïso et al., 2023).

Furthermore, there is a severe lack of field literature that statistically analyzes and interprets the moderating role of structural barriers and stresses (such as war, suspension of salaries, water and logistical poverty, and pathological pregnancy) in weakening the explanatory power of education and theoretical awareness and hindering their translation into actual practices in war-affected societies such as Yemen in 2026.

### Research Gap and Scientific Distinction of the Current Study

The current study addresses the epistemological and methodological shortcomings of previous literature and distinguishes itself through three critically important structural and scientific aspects:

•**Analytical predictive power:** The study uses a statistical regression model to measure the predictive impact of nutritional awareness, culture, and behavioral practices as independent variables on **shared maternal and fetal health and clinical indicators** (such as hemoglobin level, estimated fetal weight, fetal movement and vitality, and perceived bone and dental health) as dependent variables

•**Structural and comparative diagnosis of structural determinants:** The study does not merely describe barriers as isolated figures but subjects them to inferential statistical analysis to deconstruct how these obstacles (such as purchasing power and morning sickness) intersect with the mother’s demographic and economic characteristics, thereby elucidating the gap between awareness and behavior in conflict settings.

•**Focus on Sensitive Care and Midwifery:** By providing a conceptual framework that directly serves midwives and nurses in Sana’a, linking the quality of healthcare delivery and nursing standards to the promotion of mothers’ actual behavioral practices and the avoidance of depleting their cellular biological reserves.

## 4. Methodology

### Research Design, Study Population, Sample, and Temporal and Spatial Limitations

The study adopted a descriptive-analytical approach due to its high suitability for assessing and examining public health dimensions, the impact of human behavior, and predicting causal relationships. The study population was defined as all pregnant women attending hospitals, health centers, and major government and specialized educational clinics within the geographical scope of the Capital Secretariat of Sana’a, Republic of Yemen, during the data collection period in December 2025.

Given the inability to reach all members of the study population, a convenience sample of 200 pregnant women was selected using simple random sampling to ensure representation and equal opportunity for all demographic, social, and economic backgrounds of the participants. Strict clinical inclusion criteria were established, requiring that the mother be in the second or third trimester of pregnancy to ensure stable nutritional practices and the fetus’s development and notable physiological stability. The field sample size was set at 200 pregnant women based on strict methodological and statistical determinants that ensure the statistical power of the linear regression model used. The adequacy of this sample size (N=200) can be scientifically justified as follows:

**- Standard heuristic rules for sample size calculation in regression analysis (Heuristic Rules for Regression):** Based on established methodological literature (such as Green’s rule, 1991; and the rule by Tabachnick & Fidell, 2013), the minimum acceptable sample size for testing the overall multiple regression model is calculated using the methodological formula: N ≥ 50 + 8m (where m, representing the number of independent variables or dimensions included in the model). Given that the current study includes two main dimensions as predictive independent variables (food awareness and culture, and behavioral practices), the required minimum is 50 + 8(2) = 66 cases only. Similarly, the test for individual predictors requires a sample size calculated according to the formula: N ≥ 104 + m, i.e., 104 + 2 = 106 cases. Consequently, the current sample size (N=200) far exceeds all psychometric and statistical minimum thresholds, granting the mathematical model a high degree of freedom that prevents Type I or Type II errors.

**- Post-hoc Power Analysis:** Using the statistical power analysis software (G*Power 3.1) and applying the protocol for multiple linear regression tests (F-tests, Linear Multiple Regression: Fixed model, R² deviation from zero), and incorporating the current study parameters: a significance level of α = 0.05, a medium effect size (f^2^= 0.15 according to Cohen’s classification), and the presence of predictor variables; the analysis showed that a sample size of N = 200 provides an effective statistical power (Statistical Power: 1 -beta) exceeding the threshold of (0.95), which far exceeds the traditional international benchmark recommended in clinical and health studies, which is (0.80).

**- Structural and Logistical Determinants in Conflict Environments (Contextual & Logistical Feasibility):** In addition to statistical adequacy, the selection of a sample size limited to 200 cases complies with the criteria of realism and field feasibility in communities affected by prolonged wars and crises. The study faced highly complex security, logistical, and institutional constraints in the capital city of Sana’a due to the deterioration of hospital infrastructure and difficulties in ensuring safe access. With the application of strict clinical inclusion criteria that limited participation to pregnant women in their second and third trimesters to ensure a stable biological response, the sample of 200 cases represents the ideal and precise balance between statistical robustness and the ability to control data quality and eliminate bias in an extremely sensitive and dangerous field environment.

### Data Collection Tool and Psychometric Testing (Validity and Reliability)

The primary data collection tool was a scientifically structured and validated questionnaire consisting of 42 items, constructed and designed based on a thorough review of the existing literature and recommendations from the World Health Organization (WHO, 2024) and international partner organizations (UNFPA, 2025; UNICEF, 2024, 2025). The items in the instrument were statistically distributed to measure five main dimensions: demographic, functional, and obstetric characteristics (6 items), nutritional awareness and culture (10 items), actual dietary practices and behaviors (12 items), barriers and influencing factors (8 items), and maternal and fetal health and clinical indicators (12 items). A three-point Likert scale (Yes = 3, Sometimes = 2, No = 1) was adopted to facilitate understanding, minimize confusion among mothers, and reduce the reporting burden during the face-to-face interview.

To verify the internal consistency and reliability of the instrument’s items, *Cronbach’s alpha* coefficients and Pearson correlation matrices for the subscales were calculated, as shown in the following detailed tables:

**Table (1):**
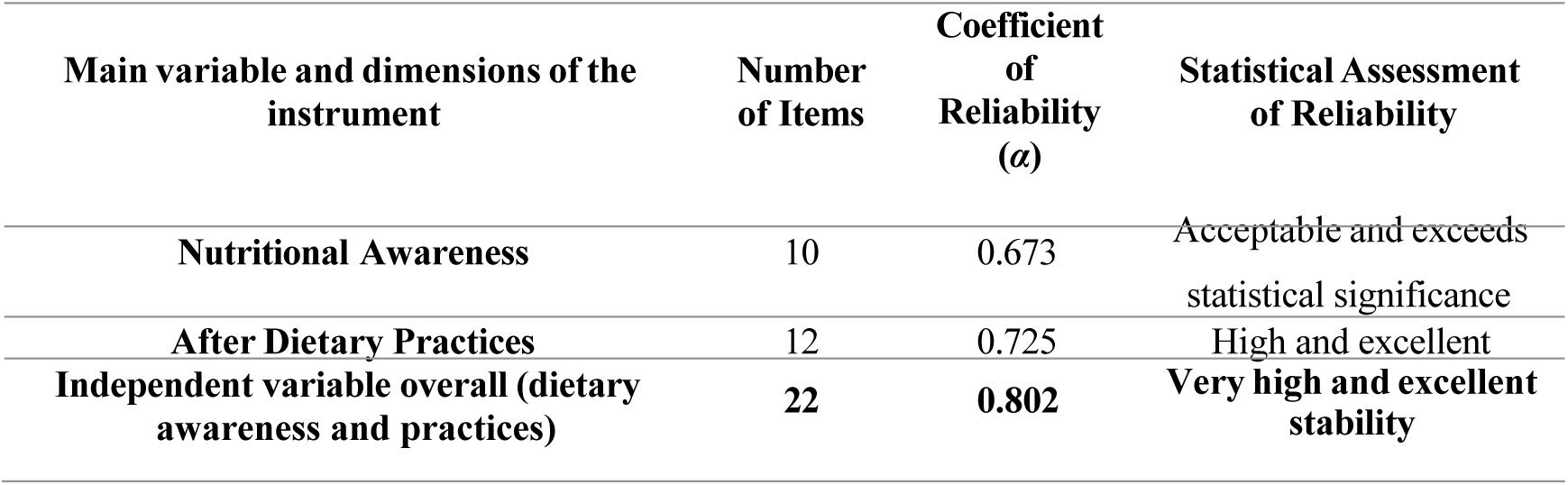
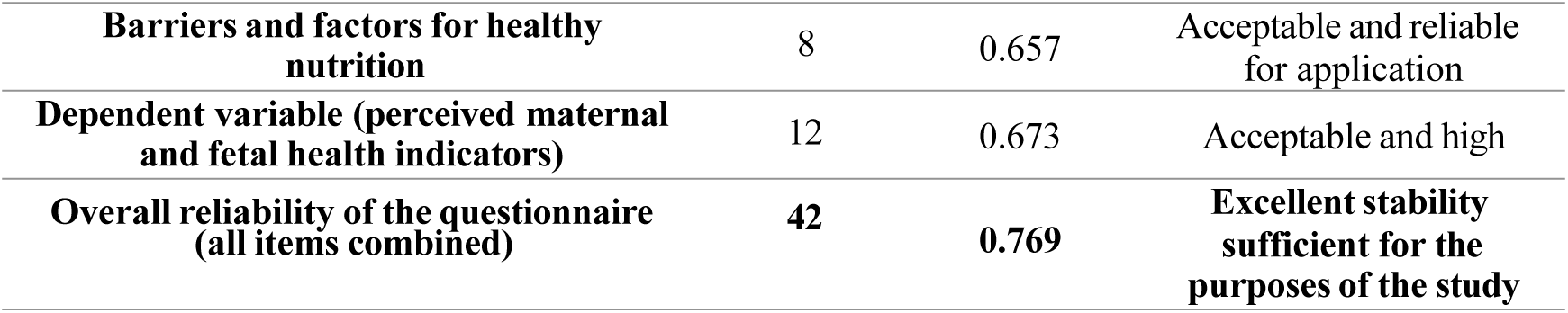
Cronbach’s Alpha Stability Coefficients for the Study’s Dimensions and Areas (n = 200)

| Main variable and dimensions of the instrument | Number of Items | Coefficient of Reliability ($\alpha$) | Statistical Assessment of Reliability |
| --- | --- | --- | --- |
| Nutritional Awareness | 10 | 0.673 | Acceptable and exceeds statistical significance |
| After Dietary Practices | 12 | 0.725 | High and excellent |
| Independent variable overall (dietary awareness and practices) | 22 | 0.802 | Very high and excellent stability |
| **Barriers and factors for healthy nutrition** | 8 | 0.657 | Acceptable and reliable for application |
| **Dependent variable (perceived maternal and fetal health indicators)** | 12 | 0.673 | Acceptable and high |
| **Overall reliability of the questionnaire (all items combined)** | **42** | **0.769** | **Excellent stability sufficient for the purposes of the study** |

The data presented in Table (1) conclusively demonstrate the quality of the psychometric properties of the instrument; all Cronbach’s alpha coefficients for the subscales exceeded the scientifically accepted threshold (0.60), ranging from (0.657 and 0.802), while the overall reliability of the instrument was 0.769, confirming the high internal consistency and statistical coherence of the questionnaire items and their absolute validity for obtaining unbiased field results.

**Table (2):** Pearson correlation coefficient matrix for the sub-dimensions of the independent variable (awareness and practices)

| **Subdimensions and Independent Variable** | **Total Awareness and Practices** | **Awareness and Food Culture** | **Dietary Practices and Behaviors** |
| --- | --- | --- | --- |
| **Total Awareness and Practices** | 1 |  |  |
| **Food Awareness and Culture** | 0.894** | 1 |  |
| **Dietary Practices and Behaviors** | 0.874 | 0.562** | 1 |

**\* The symbol (\*\*) indicates statistical significance at a highly significant level ( $p < 0.001$ )**

The Pearson correlation matrix in Table (2) shows a positive, direct, and statistically significant correlation (*p*<0.001) between the dimensions of the independent variable and among themselves, as well as with the total score; the correlation coefficient between awareness and practice was 0.562, and the correlation values for the dimensions with the total variable ranged between (0.874-0.894). This relationship confirms the construct validity and internal consistency of the instrument and the reliability of its items in measuring the cognitive and behavioral determinants among pregnant women who visit medical facilities in Sana’a.

### Data Quality Assessment and Statistical Assumptions Tests for Regression

Before proceeding to test the hypotheses and conduct impact and causal prediction tests, the field data were examined and cleaned through two main stages to ensure the model was free of mathematical flaws:

**1. Missing Values Treatment:** Initial examination of the data entered into SPSS revealed missing cells resulting from the failure to fill in some options, accounting for a very small proportion (1.21%) of the total sample cells (Figure 1 and Figure 2 before and after treatment). All these missing values were treated and statistically replaced in full using the weighted arithmetic mean method for each item, which provided a 100% complete and comprehensive database without causing any bias in the field study results.

**2. Normality Test:** Skewness and kurtosis tests were conducted for all variables to ensure the validity of parametric tests. The results showed that the skewness and kurtosis values for all sub-study axes fell entirely within the statistically acceptable range (±2.58) at the enhanced significance level (0.01), confirming that the data follow a symmetric normal distribution (Figure 9 for residuals) and allowing for the use of parametric regression and variance tests.

**3. Multicollinearity and Autocorrelation Test:** Values for the Variance Inflation Factor (VIF) and Tolerance were extracted to verify the absence of spurious multicollinearity between the dimensions of awareness and practices. The VIF values for the dimensions were 1.483 and 1.459, which are well below the statistical threshold (10), while the tolerance values were 0.675 and 0.685, which are well above the acceptable minimum (0.10). The Durbin-Watson statistic recorded a value of (1.653), which falls within the ideal independent range of (1–3), fully confirming that the regression model is free of multicollinearity and autocorrelation of errors, that the errors are randomly distributed (Scatterplot in Figure 10), and that the model is valid for scientific interpretation and generalization.

## 5. Study Results

### Descriptive analysis of the study variables and dimensions from the perspective of the research sample

The verbal estimate of the degree of agreement and the corresponding percentages were calculated based on the application of the three-point Likert scale (low, medium, high) to diagnose the reality of the variables and their dimensions and formulate a scientific interpretation of the perspectives of pregnant women visiting hospitals as follows:

### First: Assessment of the levels of the independent variable (awareness and dietary practices)

The data in Table (3) show that the overall arithmetic mean for the food awareness and culture dimension among the research sample reached (2.62) with an overall agreement and application rate of 87%, indicating a high level of awareness, which points to a distinct accumulation of knowledge and a developed food culture among pregnant women in Sana’a. Paragraph (5), regarding the importance of calcium for bone formation and protection, received the highest arithmetic mean of 2.87 and a 96% agreement rate), followed by item (10) regarding awareness of the dangers of raw foods to prevent toxoplasmosis with a mean of 2.82 and a 94% rate, and awareness of the benefits of early folic acid intake with a mean of 2.77 and a 92% rate.

**Table (3):**
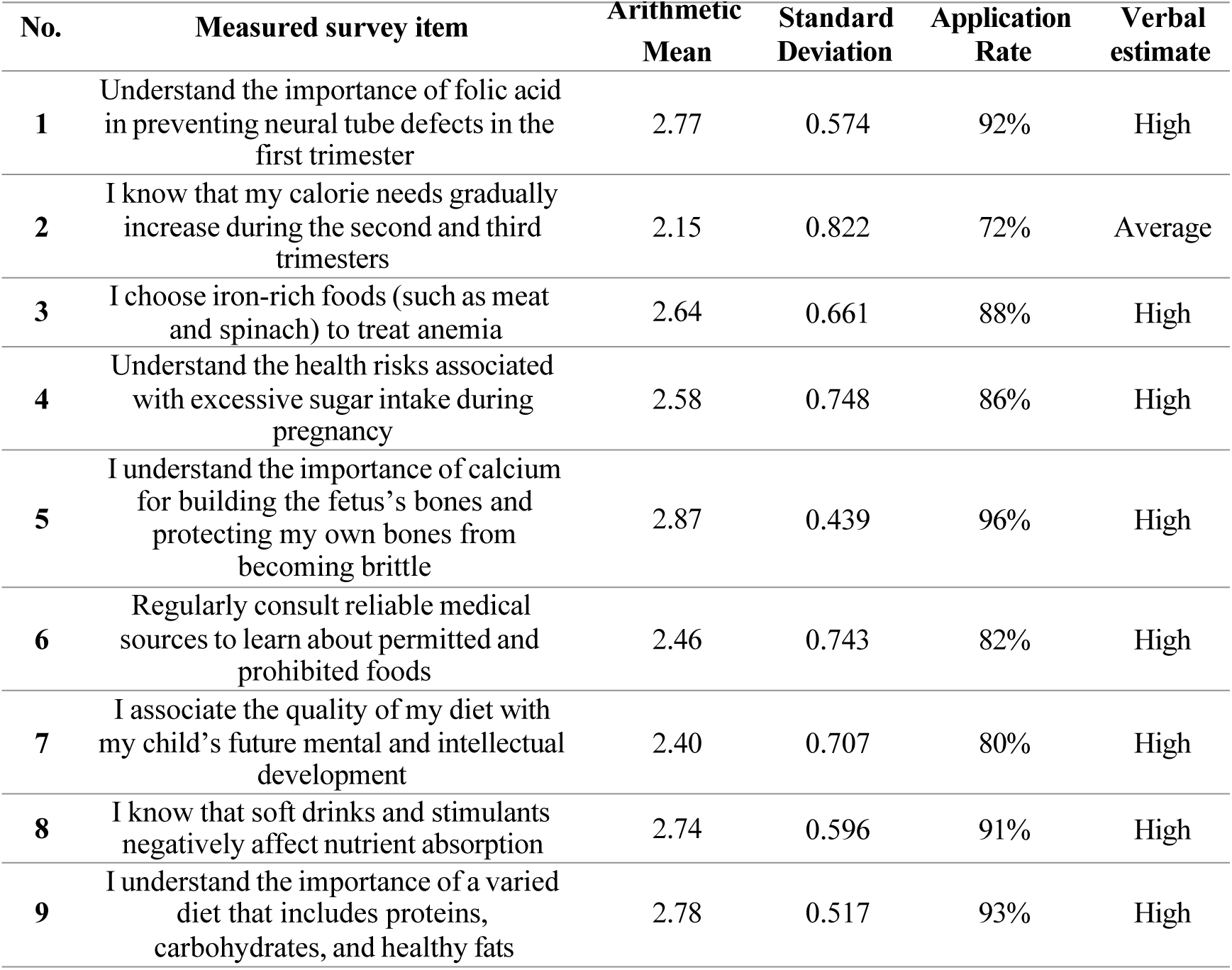
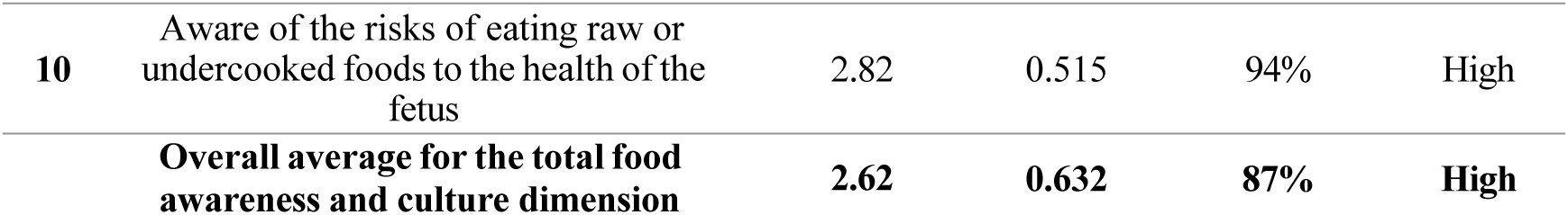
Arithmetic Means and Standard Deviations for the Level of Nutritional Awareness and Culture (n = 200)

In contrast, paragraph (2), concerning knowledge of the gradual increase in caloric and energy needs during the second and third trimesters of pregnancy, recorded the lowest arithmetic mean in the dimension at (2.15 and 72%), with a moderate level of agreement, revealing a specialized knowledge gap among mothers regarding the calculation and balance of the daily caloric intake required for rapid fetal growth.

The results in Table (4) reveal that the overall mean for the actual dietary practices and behaviors dimension recorded a high value of 2.39, with an 80% rate of agreement and field application. Preventive behavioral practices aimed at avoiding microbial and parasitic infections received the highest average scores for full agreement; paragraph (15), “Cook meat and eggs thoroughly,” recorded a mean of 2.95 and a 98% implementation rate, followed by item (21)—washing vegetables and fruits thoroughly before consumption—with an average of 2.93 and a 98% implementation rate, reflecting extremely high levels of adherence that indicate an innate desire and a strong preventive commitment to protecting the fetus from prevalent epidemics.

**Table (4):**
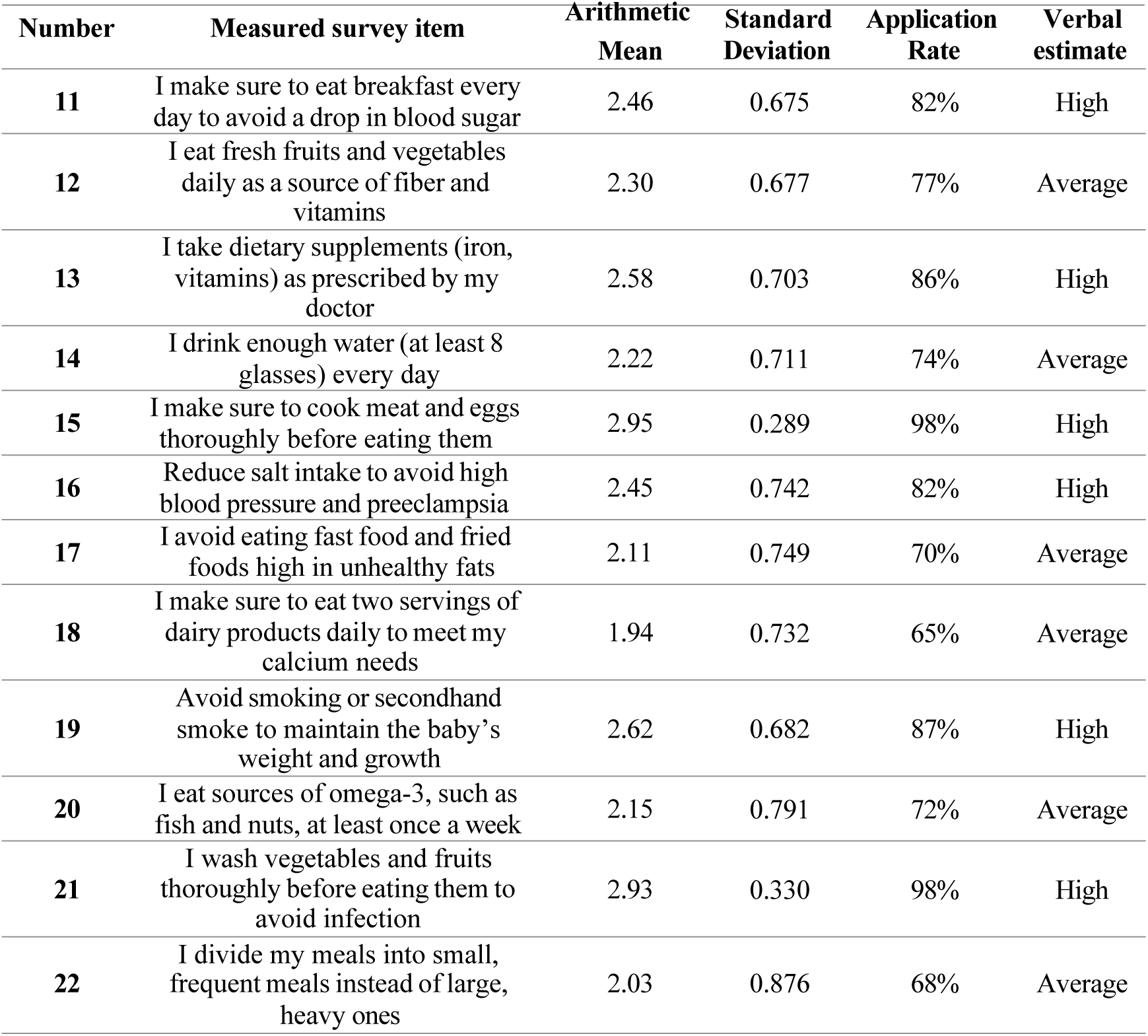
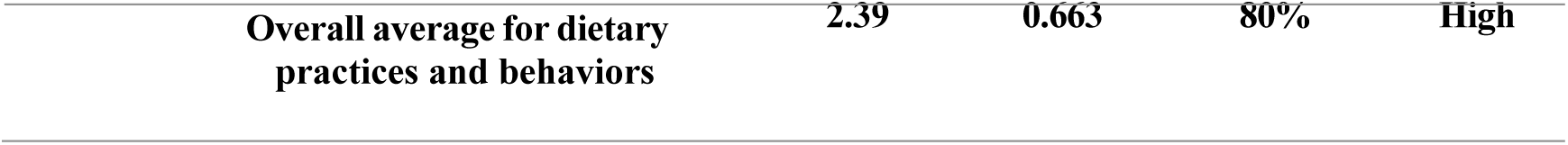
Arithmetic Means and Standard Deviations for the Level of Adherence to Dietary Practices and Behaviors (n = 200)

In contrast, behaviors related to dietary diversity and dairy consumption declined, reaching the lowest average score in the dimension for paragraph (18): “I make sure to consume two servings of dairy,” with an average of 1.94 and a compliance rate of 65%, followed by the practice of dividing meals to avoid indigestion, with an average of 2.03 and a rate of 68%, at moderate levels of agreement. This reflects the direct impact of low monthly income on limiting the financial ability to obtain the prescribed daily portions of milk products and sufficient protein.

### Second: Evaluation of the variable “obstacles and influencing factors” as a model adjustment variable

The data in Table (5) show that the overall assessment of the presence of barriers in the lives of pregnant women in Sana’a stands at moderate levels overall, with a general arithmetic mean of 1.86 and a prevalence and availability rate of 62%. The section on the impact of morning sickness and nausea on healthy eating recorded the highest arithmetic mean (2.39, with an agreement rate of 80%) at a high level, which fully supports the severe hindering effect of the physiological condition of the pregnant woman in undermining her appetite and aspirations for sound health.

**Table (5):** Arithmetic Means and Standard Deviations for the Level of Barriers and Factors Affecting Proper Nutrition (n = 200)

| No. | Measured survey item | Arithmetic Mean | Standard Deviation | Application Rate | Verbal estimate |
| --- | --- | --- | --- | --- | --- |
| 23 | The economic situation (high prices) prevents me from buying some necessary healthy foods | 2.12 | 0.755 | 71% | Average |
| 24 | I find it difficult to prepare healthy meals due to time constraints or work circumstances | 1.78 | 0.849 | 59% | Average |
| 25 | Nausea and morning sickness significantly affect my ability to eat healthy | 2.39 | 0.743 | 80% | High |
| 26 | I lack support and encouragement from my spouse or family to follow a healthy diet | 1.81 | 0.871 | 60% | Average |
| 27 | I have difficulty clearly understanding the instructions of the doctor or dietitian | 1.55 | 0.757 | 52% | Low |
| 28 | Social customs and traditions influence the type of food I eat | 1.72 | 0.844 | 57% | Average |
| 29 | I face a shortage of healthy food products in my residential area | 1.86 | 0.833 | 62% | Average |
| 30 | I rely on advice from relatives and friends regarding my diet more than on medical sources | 1.63 | 0.810 | 54% | Low |
| **Overall average for the variable "Barriers and Influencing Factors"** |  | **1.86** | **0.808** | **62%** | **Average** |

Financial constraints linked to declining purchasing power ranked second, with an arithmetic mean of 2.12 and a 71% agreement rate, as a structural barrier preventing the diversification of the mother’s food basket. The item regarding following the traditional advice of relatives and grandmothers scored low, with an average of 1.63 and a 54% agreement rate, confirming that contemporary societal trust in official medical consultations and sources has increased compared to advice from the traditional social environment.

### Third: Assessment of the Maternal and Fetal Health Indicators Variable (Combined Clinical Outcomes)

The data in Table (6) reveal that the overall health status of the mother and fetus falls within the scientifically acceptable range, with a total arithmetic mean of 2.24 and an overall agreement rate of 75%, indicating an average level of availability. The section on the absence of major metabolic and vascular disorders (preeclampsia and gestational diabetes) in the current pregnancy recorded the highest arithmetic mean, with a value of 2.71 and a 90% agreement rate. Regular fetal movement within the uterus and standard growth corresponding to gestational age as determined by ultrasound received very high agreement scores of 2.56 and 2.35, with availability rates of 85% and 78%, respectively, demonstrating the stability of the fetuses’ biological development and growth.

**Table (6):** Arithmetic Means and Standard Deviations for the Availability Level of Maternal and Fetal Health Indicators (n = 200)

| No. | Survey item measuring clinical outcomes | Arithmetic Mean | Standard Deviation | Application Rate | Verbal Rating |
| --- | --- | --- | --- | --- | --- |
| 31 | I feel energetic and lively and go about my daily life without much strain | 1.80 | 0.667 | 60% | Average |
| 32 | Medical test results show that my hemoglobin level is within the normal range | 2.56 | 0.742 | 85% | High |
| 33 | The fetus is growing normally and its weight is appropriate for the gestational age according to the ultrasound | 2.35 | 0.896 | 78% | High |
| 34 | I suffer from frequent dizziness or headaches (reverse scale measuring anemia and blood pressure) | 2.20 | 0.729 | 73% | Average |
| 35 | I am keeping my weight gain within the normal recommended range for pregnant women | 2.21 | 0.801 | 74% | Average |
| 36 | I have a strong immune system and do not frequently contract infectious diseases | 2.26 | 0.743 | 75% | Average |
| 37 | I feel the baby move regularly and with good strength | 2.56 | 0.606 | 85% | High |
| 38 | My current pregnancy has been free of gestational diabetes or high blood pressure so far | 2.71 | 0.691 | 90% | High |
| 39 | I notice that my teeth and bones are healthy and that there is no abnormal pain in them | 2.10 | 0.906 | 70% | Average |
| 40 | I enjoy a stable mental state, free from the severe mood swings associated with malnutrition | 1.84 | 0.761 | 61% | Average |
| 41 | I sleep well and do not suffer from severe insomnia at night | 2.02 | 0.797 | 67% | Average |
| 42 | My body recovers quickly from any sudden health issues during pregnancy | 2.21 | 0.725 | 74% | Average |
| **Overall average for the total maternal and fetal health indicators variable** |  | **2.24** | **0.755** | **75%** | **Average** |

In contrast, there is a clear decline and sharp drop in the mother’s physical, psychological, and clinical indicators; the section on feelings of activity, vitality, and ability to exert effort recorded the lowest arithmetic mean in the dimension at 1.80 and a 60% agreement rate, followed by a decline in levels of psychological stability, mood balance, and exposure to insomnia at low rates (1.84 and 2.02, with agreement rates of 61% and 67%, respectively), clinically revealing a decline in the pregnant mother’s energy and vitality to compensate for and meet the physiological and biological requirements necessary for healthy fetal growth.

### Fourth: Structural Summary of the Main Study Variables and Their Ranking

Table (7) confirms that the mother’s awareness and theoretical knowledge rank first as the highest field availability at 87%, significantly outperforming the actual behavioral dimension of application and practices, which ranked second at 80%. This statistically supports the formulation of a theoretical behavioral model based on the widening gap between awareness and behavior under the pressure of complex material realities.

**Table (7):** Arithmetic means, standard deviations, and percentages of the study variables and their overall dimensions.

| **Rank** | **Variable / Main Dimension Measured** | **Arithmetic Mean** | **Standard Deviation** | **Availability / Application Rate** | **Verbal assessment** |
| --- | --- | --- | --- | --- | --- |
| **1** | After Nutritional Awareness | 2.62 | 0.632 | 87% | High |
| **2** | Dietary Practices | 2.39 | 0.663 | 80% | High |
|  | **Independent variable as a whole (overall awareness and practices)** | **2.51** | **0.650** | **84%** | **High** |
| **4** | Barriers and factors affecting proper nutrition | 1.86 | 0.808 | 62% | Average |
| **3** | Dependent variable (perceived maternal and fetal health indicators) | 2.24 | 0.755 | 75% | Average |

### Hypothesis Testing, Predictive Power, and Causal Impact of the Regression Model

To verify the hypotheses and formulate the mathematical prediction and causal modeling of the effect of the total independent variable and its sub-dimensions on the dependent variable, simple and multiple linear regression tests were conducted as follows:

### First: Testing the First Main Hypothesis (The Effect of Overall Awareness and Practices on Maternal and Fetal Health Indicators)

Simple linear regression analysis was applied to measure the direct effect of the overall independent variable (overall awareness and practices) on the dependent variable (health indicators for the mother and fetus), and the results are presented in the following table:

**Table (8):**
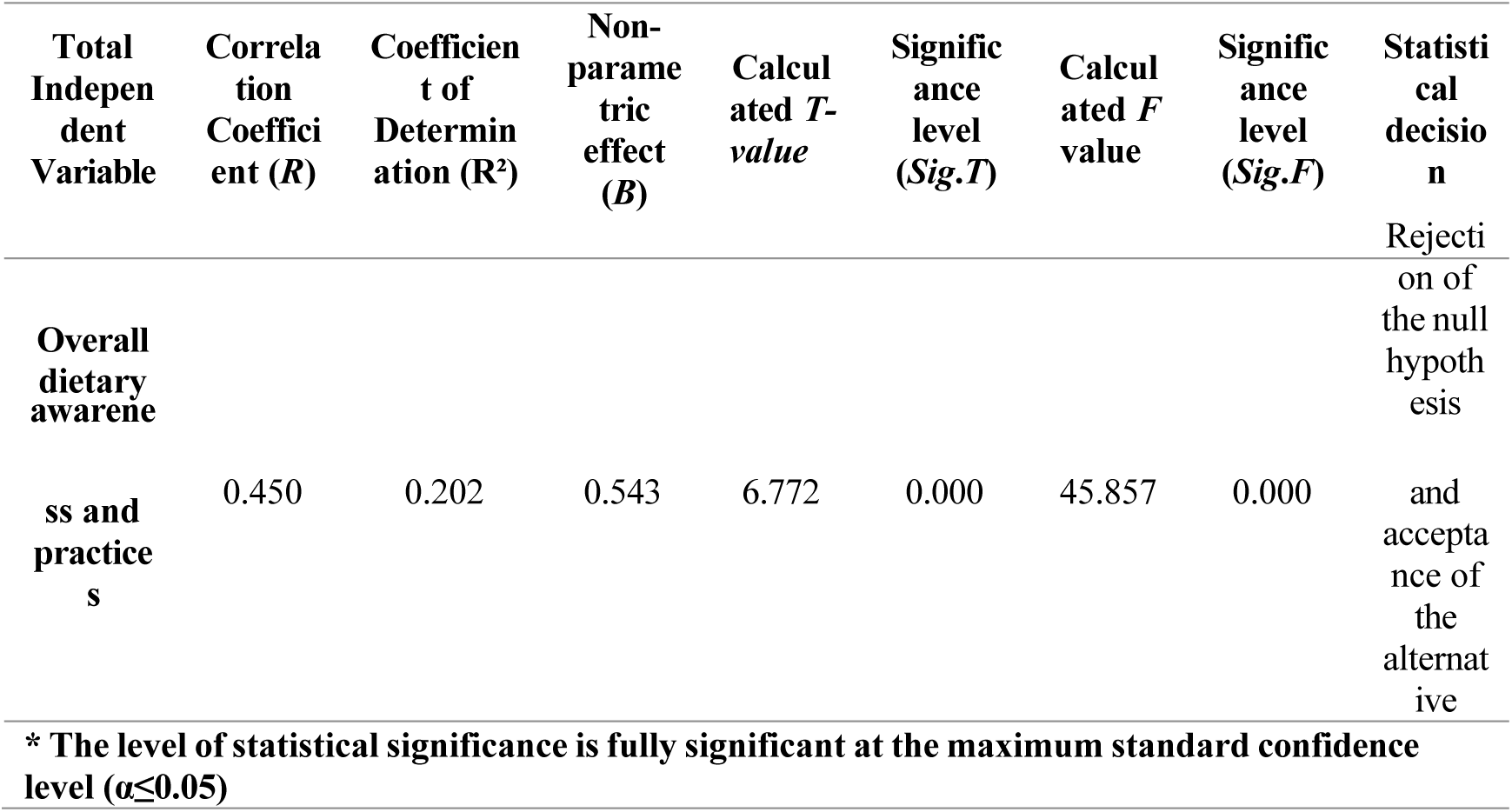
Results of the simple linear regression test of the effect of overall awareness and practices on improving maternal and fetal health indicators.

The precise scientific data in Table (8) confirm the existence of a positive, moderately strong linear correlation with a value of (R=0.450), which is statistically significant based *on the F-test* for the regression model (45.857) at a highly significant level (*p*<0.001). The coefficient of determination was (R²= 0.202), which mathematically explains the following: **20.2% of the total variation in maternal and fetal health indicators is directly explained and controlled by the mother’s level of awareness and dietary practices**, while the remaining 79.8% is attributed to other environmental, clinical, and random variables outside the scope of the current research model.

The non-standard regression coefficient (B=0.543) implies that a 100% increase in the pregnant mother’s awareness and dietary practices would result in a direct proportional improvement of 54.3% in her health indicators and fetal growth indicators, assuming all other variables remain constant, which is a highly significant predictive finding that fully supports the rejection of the null hypothesis and the acceptance of the first alternative hypothesis.

### Second: The combined and relative effects of the dimensions of awareness and practices (multiple regression analysis)

To determine the competitive predictive power of the dimensions and identify which of the two dimensions has the decisive physiological and behavioral impact, multiple linear regression analysis was applied to model the effect, as shown in the following table:

**Table (9):**
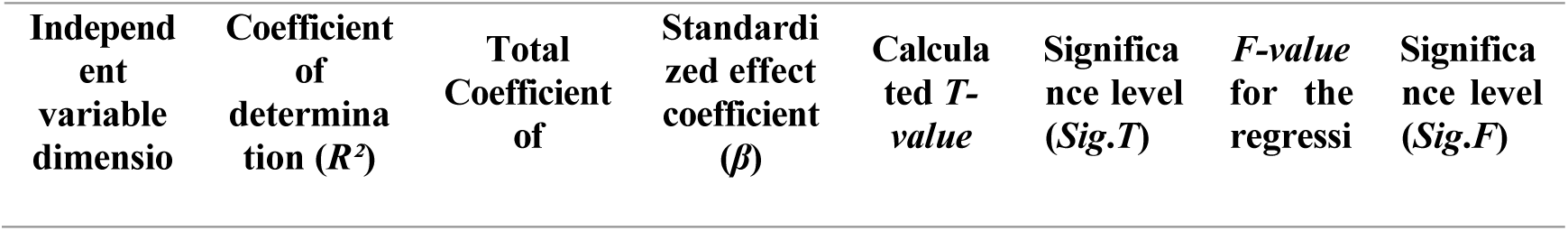
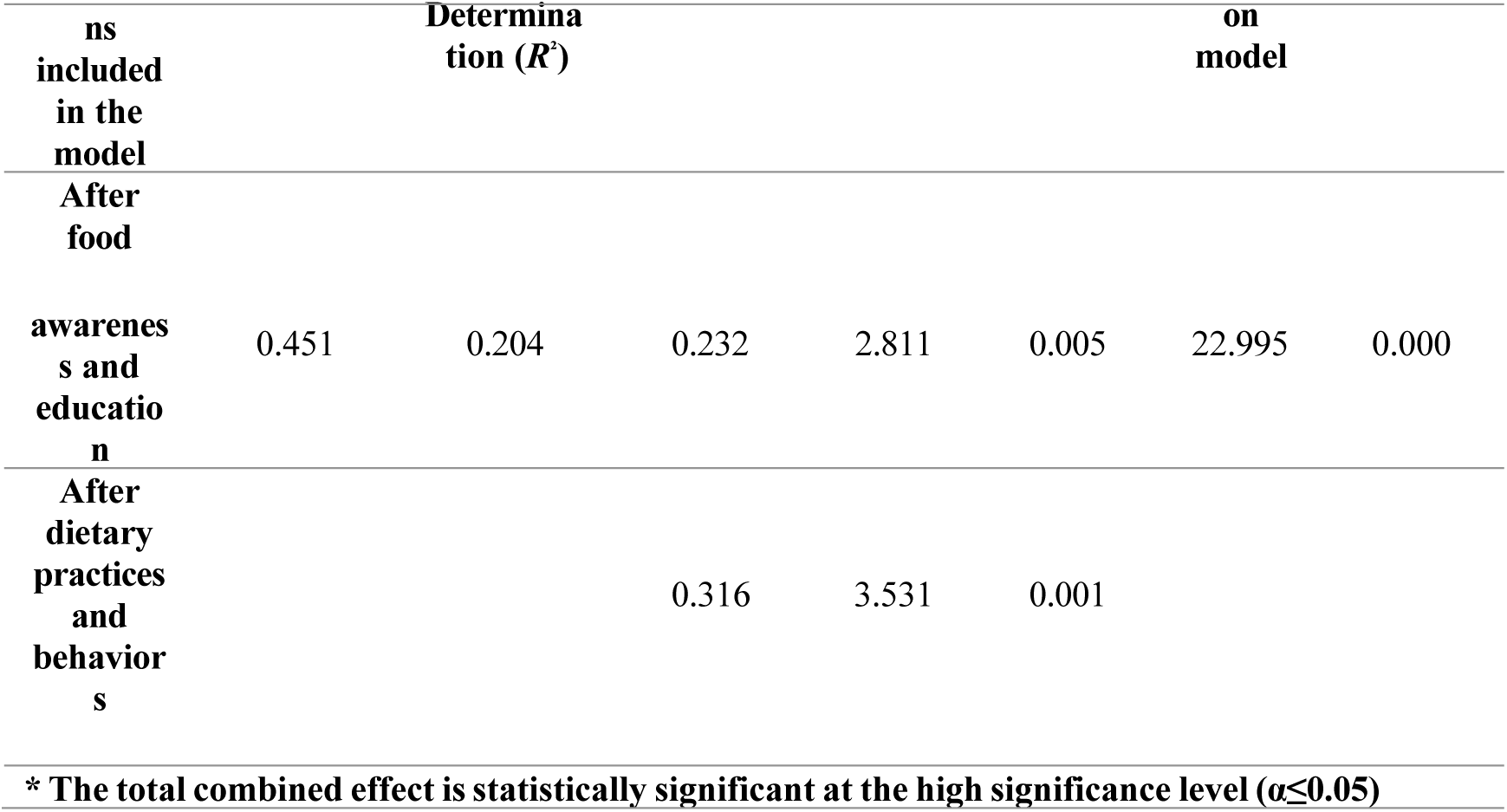
Results of the multiple linear regression test for the combined effect of the awareness and practices dimensions on maternal and fetal health indicators.

A careful reading and comparison of the multiple regression results in Table (9) reveal a highly significant statistical and structural finding; the total combined explanation for the two dimensions rose to (R²= 0.204,with an absolute statistical significance for the total regression F = 22.995). The standardized effect sizes confirm a statistically significant and meaningful advantage in favor of the actual applied behavioral dimension; the beta value for dietary practices and behaviors reached (β = 0.316,T=3.531, p=0.001), surpassing the predictive power of the theoretical awareness and culture dimension, which stood at (β=0.232, T=2.811, p=0.005).

This rigorous statistical result demonstrates that **behavioral practice and the daily physical application of healthy eating and nutrient sources represent the strongest determinant, predictor, and indicator of the health indicators of the mother and fetus compared to merely possessing theoretical cognitive awareness**, which strongly supports the hypothesis of a widening knowledge-behavior gap and the need for healthcare programs to gradually shift toward empowerment and the practical modeling of dietary behavior for pregnant women to overcome structural barriers.

### Third: Individual analysis of the impact of each dimension of awareness and practices on maternal and fetal health indicators

For further verification and testing of the sub-hypotheses, a separate simple linear regression model was run for each dimension as follows:

**Table (10):** Results of the simple linear regression test for the effect of awareness and dietary culture individually on maternal and fetal health indicators.

| Indepen<br>dent<br>variable | Coefficie<br>nt of<br>Determin<br>ation ($R^2$) | Coefficie<br>nt of<br>Determin<br>ation ($R^2$) | Direct<br>Predic<br>tive<br>Effect<br>($B$) | Calcul<br>ated $T$-<br>value | Signific<br>ance<br>level<br>($Sig.T$) | Calcul<br>ated $F$-<br>value | Signific<br>ance<br>Level<br>($Sig.F$) | Statisti<br>cal<br>decisio<br>n made |
| --- | --- | --- | --- | --- | --- | --- | --- | --- |
| Nutritio<br>nal<br>Awaren<br>ess and<br>Culture | 0.388 | 0.150 | 0.396 | 5.688 | 0.000 | 32.354 | 0.000 | Delta<br>and<br>Accept<br>ance of<br>the<br>First<br>Sub-Hypoth<br>esis |

The data in Table (10) show that cognitive awareness alone accounts for 15% of the variance in health outcomes, confirming the intrinsic value of nutritional awareness and its importance as a foundational element in shaping public health.

**Table (11):** Results of the simple linear regression test for the effect of practices and behaviors individually on maternal and fetal health indicators.

| Independent variable | Correlation Coefficient (R) | Coefficient of Determination (R<sup>2</sup>) | Direct predictive effect (B) | Calculated T-value | Significance level (Sig.T) | Calculated F-value | Significance level (Sig.F) | Statistical decision |
| --- | --- | --- | --- | --- | --- | --- | --- | --- |
| Dietary practices and behaviors | 0.411 | 0.169 | 0.457 | 6.057 | 0.000 | 36.688 | 0.000 | Delta and acceptance of the second alternative hypothesis |

The data in Table (11) clearly support the predictive gap; as the actual behavioral dimension alone accounts for 16.9% of the variance in health indicators, surpassing the cognitive explanation by nearly two percentage points on its own—a clear causal indication demonstrating the superiority of applied behavior in shaping clinical reality.

### Testing for statistical differences according to demographic, functional, and generational variables

To verify the levels of significant variance in the responses of sample participants according to differences in personal, occupational, and obstetric characteristics, the independent samples t-test and *one-way* ANOVA were applied. The results revealed the following findings:

### First: Statistical differences based on the variable of the pregnant mother’s occupational status (occupation)

*A t-test* was applied to compare the means of housewives and employed women, and the results are presented in the following table:

**Table (12):**
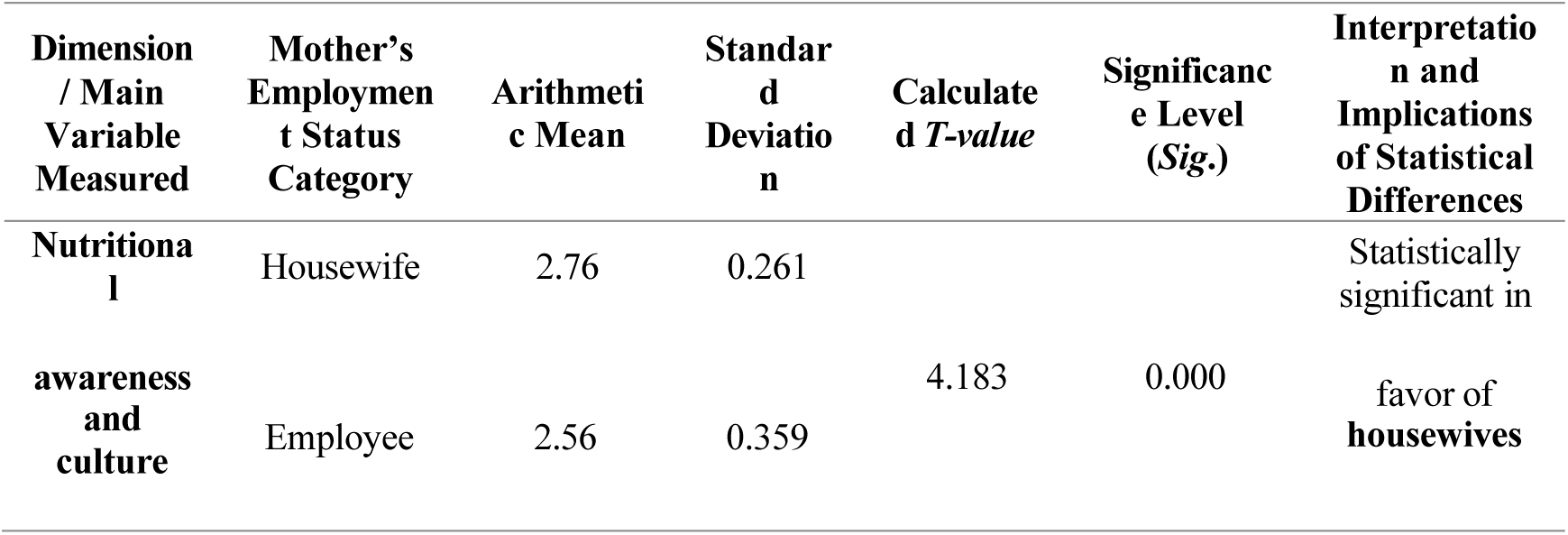
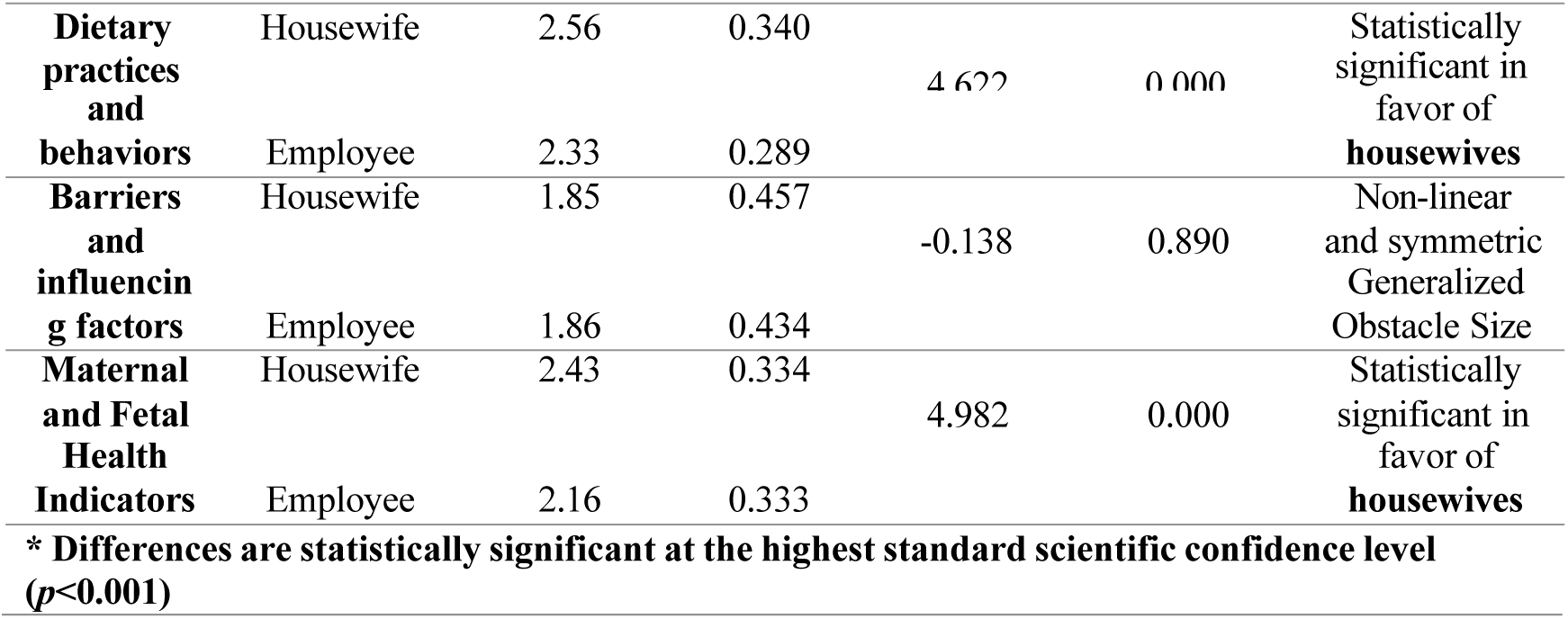
Results of the statistical difference test for study variables based on the mother’s employment status (housewife, employee)

The scientific data in Table (12) confirm the existence of highly statistically significant differences in awareness, practices, and health indicators for the mother and fetus in favor of housewives; their average ratings of the reality of implementation and health status (2.76, 2.56, and 2.43, respectively) significantly exceeded those of employed women (2.56, 2.33, and 2.16, respectively). This decline in behavioral and health outcomes among employed women is physiologically and practically attributed to the dual burden and stress faced by working women in Sana’a, as well as the limited time available to prepare fresh, healthy meals and adhere to regular checkups and nutritional monitoring, while the two groups show no differences in constraints due to the parity and equality of their general living conditions under the pressures of conflict and shared poverty.

### Second: Statistical differences according to the age group variable of pregnant women

A one-way ANOVA was applied to compare the four age groups, and the results are presented in the following table:

**Table (13):**
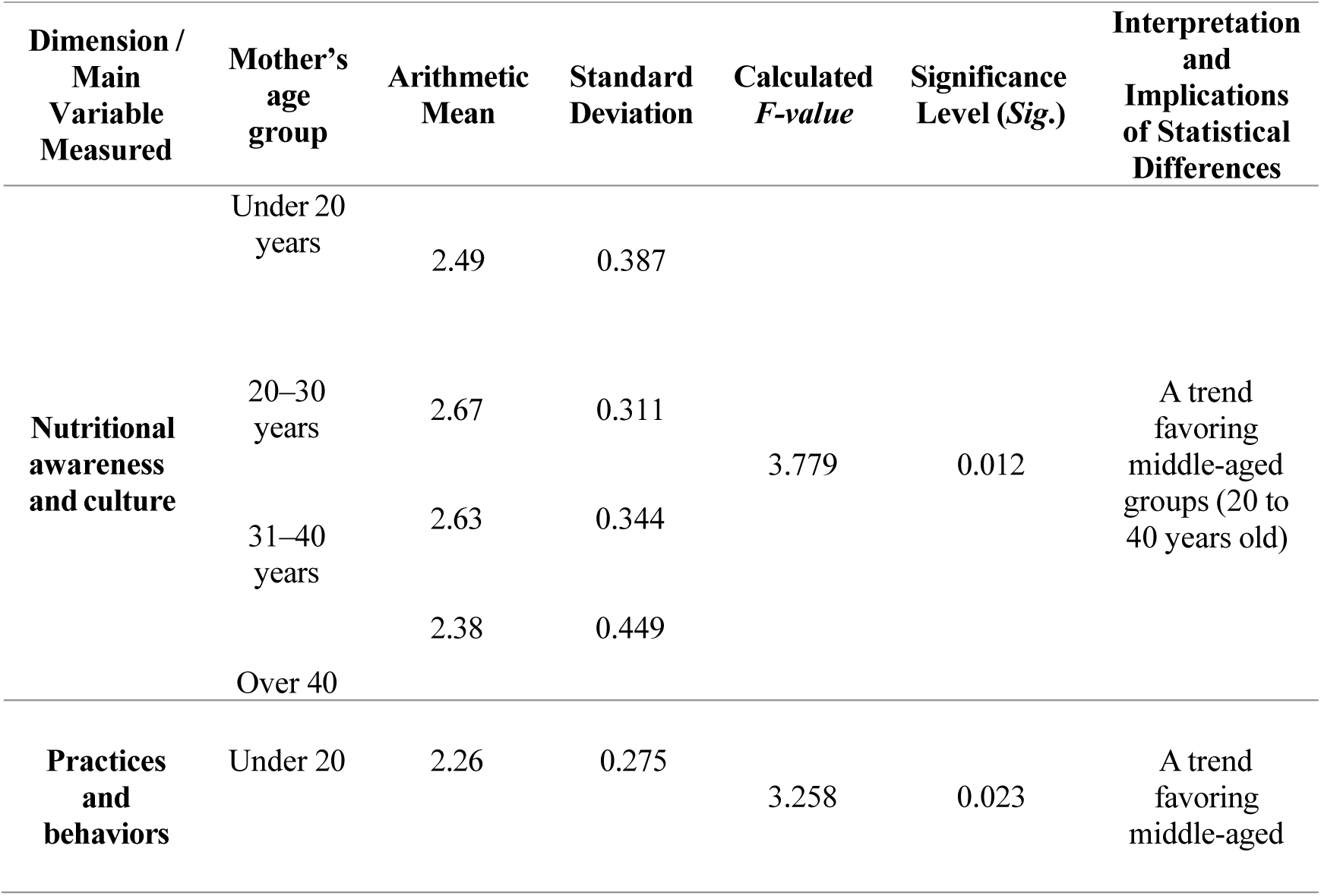
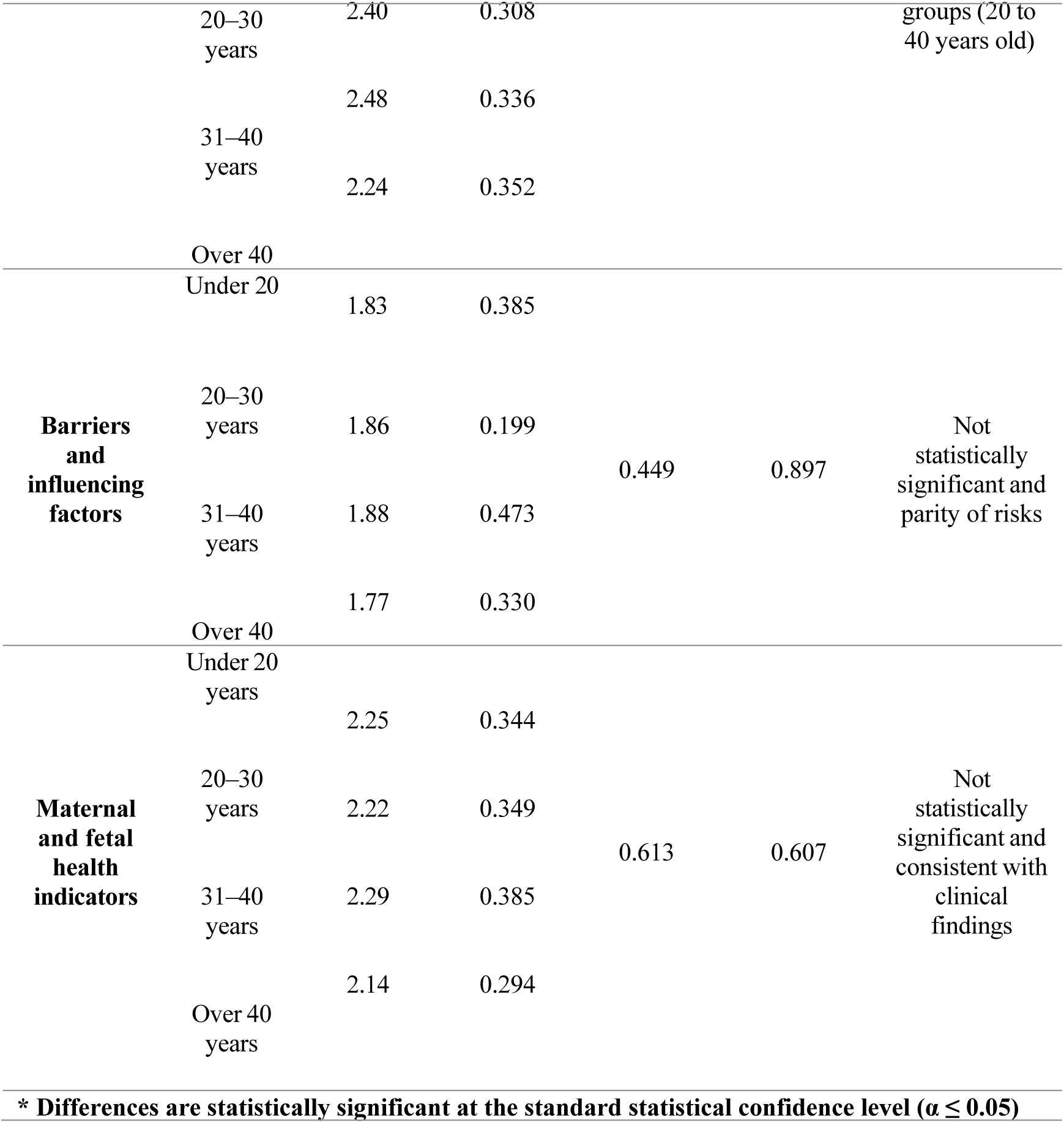
Results of the test of differences for study variables according to the age group of the pregnant mother (n = 200)

The data in Table (13) demonstrate statistically significant differences in the dimensions of awareness and dietary practices in favor of the middle age groups (20 to 40 years). This result is statistically explained by the fact that this age group represents the ideal physiological combination of full cognitive and perceptual maturity, physiological fertility, and social stability for the pregnant mother. In contrast, younger mothers (under 20 years old) lagged behind due to their limited cognitive and behavioral experience and physical maturity, while older mothers (over 40 years old) lagged behind due to a decline in their cognitive interest or adherence to traditional, culturally entrenched behavioral patterns.

### Third: Statistical Differences According to the Educational Level of the Pregnant Mother

A one-way ANOVA was applied to compare educational levels, and the results are presented in the following table:

**Table (14):**
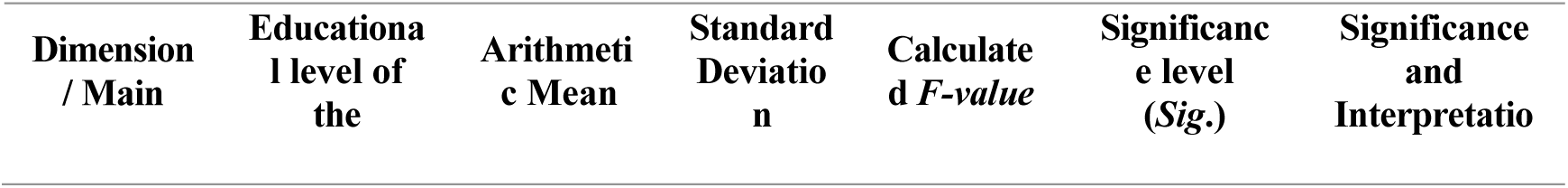
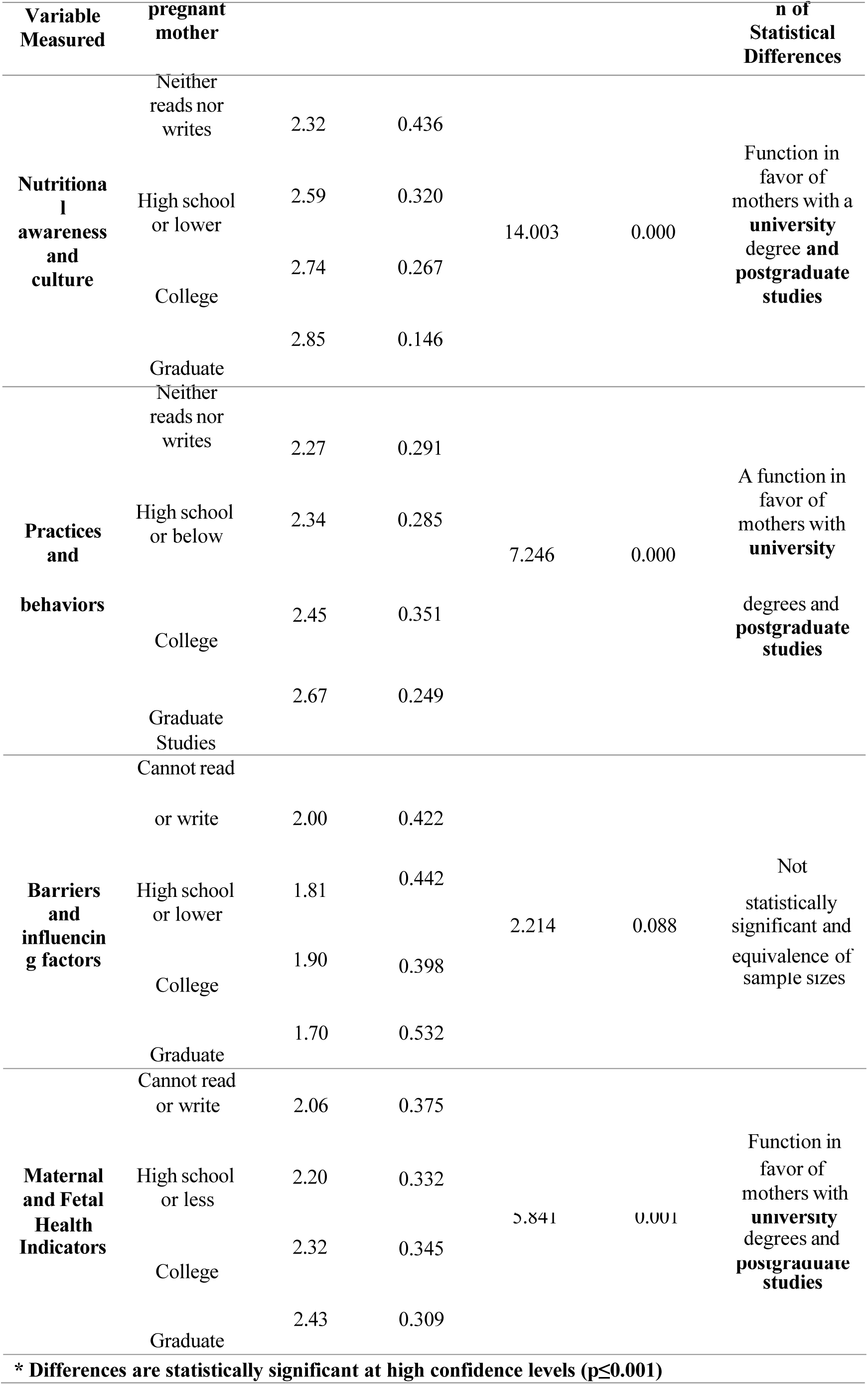
Results of the test of differences for study variables based on the mother’s educational level (n = 200)

The results in Table (14) confirm the decisive and prominent role of education and intellectual empowerment in shaping nutritional and health outcomes; there are highly significant differences in awareness, behavioral practices, and health indicators for both mother and fetus in favor of the university and postgraduate education groups. Mothers with postgraduate education recorded the highest cognitive, behavioral, and health scores on the scale (2.85, 2.67, and 2.43, respectively). This statistical finding confirms that a mother’s intellectual capital endows her with advanced cognitive skills to interpret medical information, seek affordable and accessible dietary alternatives, and address resource constraints, resulting in superior health outcomes and a favorable biological response.

### Fourth: Statistical Differences According to the Variable of Monthly Household Income

A one-way analysis of variance was applied to compare income categories, and the results are presented in the following table:

**Table (15):** Results of the test of differences for study variables based on the monthly household income variable (n = 200)

| **Dimension / Main variable measured** | **Monthly household income category** | **Arithmetic Mean** | **Standard Deviation** | **Calculated *F*-value** | **Significance Level (*Sig.*)** | **Significance and Direction of Statistical Differences** |
| --- | --- | --- | --- | --- | --- | --- |
| **Nutritional awareness and culture** | Low | 2.45 | 0.394 | 6.169 | 0.003 | A measure that benefits middle- and high-income groups |
|  | Medium | 2.66 | 0.316 |  |  |  |
|  | High | 2.69 | 0.345 |  |  |  |
| **Practices and behaviors** | Low | 2.24 | 0.269 | 7.389 | 0.001 | Favors middle- and high-income groups |
|  | Medium | 2.41 | 0.307 |  |  |  |
|  | High | 2.54 | 0.375 |  |  |  |
| **Obstacles influencing factors** | Low | 2.09 | 0.493 | 12.605 | 0.000 | Function in favor of low-income individuals (most in need and facing obstacles) |
|  | Medium | 1.84 | 0.409 |  |  |  |
|  | High | 1.55 | 0.268 |  |  |  |
| **Maternal and Fetal Health Indicators** | Low | 2.11 | 0.377 | 5.909 | 0.003 | Favors middle- and high-income groups |
|  | Medium | 2.24 | 0.342 |  |  |  |
|  | High | 2.42 | 0.300 |  |  |  |

**\* Differences are statistically highly significant at the standard confidence level ( $p \leq 0.003$ )**

The detailed statistical data presented in Table (15) demonstrate the dominance and influence of a mother’s economic status and capacity on her dietary choices and biological indicators; there are statistically significant differences in awareness, practices, and health indicators in favor of those with high and middle incomes. In contrast, low-income mothers outperformed their counterparts in confronting and recognizing barriers at very high and statistically significant rates, with a mean of (2.09), compared to high-income mothers, who recorded the lowest rate of barriers at (1.55). This result confirms that poverty and low financial capacity represent the strongest structural barrier and determinant that undermines the effectiveness of individual awareness and prevents mothers from adopting healthy and varied dietary behaviors in war-torn environments affected by the high cost of living.

### Fifth: Statistical Differences According to Obstetric Variables (Stage of Pregnancy and Current Order)

A one-way ANOVA was applied to compare groups based on stage of pregnancy and current trimester, and the results reveal the following:

· Mother’s current stage of pregnancy: The data showed no statistically significant differences in awareness levels (p = 0.542), practices (p = 0.094), barriers (p = 0.075), or maternal and fetal health indicators (p = 0.117); as the statistical means for all pregnancy stages were very close and highly consistent.

· Order of the current pregnancy: The data showed no statistically significant differences in awareness (p = 0.427), practices (p = 0.889), barriers (p = 0.384), or health indicators (p = 0.479); as the averages for primiparous mothers were identical and similar to those of multiparous mothers up to the fifth pregnancy and beyond.

These field findings confirm an exceptional scientific fact: a mother’s experience and the repetition of previous pregnancies and births do not automatically lead to accumulated knowledge or improved dietary behavior, thereby refuting the prevailing belief that repeated pregnancies foster self-learning, and highlights the urgent need to provide guidance and training to all pregnant women equally through supported midwifery education programs, without relying on their previous experiences.

## 6. Discussion

Inferential statistical analysis of the field study data reveals a high level of nutritional awareness (87%) corresponding to an 80% implementation of nutritional practices, with a statistically significant effect of awareness and practices in explaining 20.2% of the variance in maternal and fetal health outcomes and indicators (R² = 0.202, *p* < 0.001). The relative strength of the effect in the multiple regression analysis demonstrates that the predictive power of actual behavioral practices (β=0.316) exceeds that of theoretical cognitive awareness (β=0.232).

This gap between guidance and biology can be analyzed and explained by drawing on the philosophical framework of “social construction theory” and the sociological reality of the Sana’a Capital Secretariat; for it is not merely the presence or absence of a mother’s nutritional awareness that constitutes the sole obstacle to sound feeding practices, but rather, the desires and cognitive constraints of pregnant women clash with suffocating material and structural pressures and barriers imposed by the conditions of conflict, poverty, and the general collapse of livelihoods (Haza’a, 2022).

Pregnant women in Sana’a, despite their full theoretical understanding of the benefits of calcium for their own protection and that of their fetus’s bones—with a high awareness rate of 96%—are behaviorally and financially unable to practice and consume daily servings of milk and dairy products, with an average that has dropped to the lowest levels of behavioral practice (1.94 and 65%), due to the deterioration of monthly household income, the skyrocketing prices, and the scarcity of financial resources, which 71% of mothers identified as a direct economic barrier to proper nutrition. This interpretive finding is fully consistent with the results of (Alsaeedi & Al-Haddad, 2026), which identified economic poverty as a decisive barrier to mothers’ access to care and education services.

### Clinical and Physiological Explanation of the Paradox Between Subjective and Objective Indicators

The findings observed in this study provide conclusive statistical and biological evidence of what can be termed the paradox of intrauterine physiological adaptation in crisis environments. Although there is a real gap between the mother’s awareness and her actual practices due to economic inflation and declining purchasing power, the measurement of outcomes showed a proportional increase and acceptable stability in fetal growth, estimated weight, and movement at rates of 78% and 85%, compared to a sharp decline in the mother’s energy and physical vitality (60%) and a deterioration in her psychological stability (61%).

This paradox finds its precise clinical explanation in the characteristics of “fetal adaptation and the pathophysiology of pregnancy,” based on the hypotheses of “maternal-fetal resource allocation conflict” from the perspective of conflict medicine and the pathophysiology of pregnancy; for within the uterus, the fetus represents a “distinct biological drain” possessing superior physiological and muscular capacity to draw and absorb essential nutrients, gases, and minerals (such as calcium, iron, and trace minerals) from the mother’s bloodstream and cellular stores to meet the demands of its rapid neurological and physical growth and development, regardless of the adequacy or deficiency of the mother’s external nutritional intake (Abu-Saad & Fraser, 2010).

Amid the complex conflict conditions in Sana’a, where mothers suffer from severe deficiencies in both micronutrients and macronutrients due to the collapse of livelihoods, an evolutionary physiological protection system is activated that grants absolute biological priority to the survival of the species (i.e., the fetus) to protect it from disruptions in early fetal programming. At that point, the mother’s body undergoes a process of “forced and systematic mobilization of maternal cellular and mineral stores” (Mobilization of Maternal Cellular and Mineral Stores); in which bone and tooth resorption is activated to release calcium, and iron and carbohydrate stores are drawn from the mother’s liver and cellular tissues (such as ferritin), and pumped through the umbilical cord to meet the fetal nervous and physical growth needs.

This intense biological draw clinically explains the high prevalence of complaints among mothers regarding bone and tooth pain (70%) and feelings of chronic fatigue and dizziness (73%) (Esquivel, 2021). This finding also provides a stark physiological explanation for the findings of a study (Alsebaeai et al., 2025) regarding the prevalence of pica and the consumption of clay and ash (28.7%) among pregnant women in Sana’a; as this behavioral disorder is nothing more than an unconscious pathophysiological manifestation triggered by the mother’s brain as a result of severe anemia and zinc and iron deficiency caused by the “biological depletion and predation” exerted by the fetus on her cellular stores under the strain of material and livelihood-related food deprivation.

### The Role of Management and Health Quality in Bridging Care Gaps

Statistical results did not demonstrate significant differences in the perception and extent of general barriers faced by housewives and working women; rather, they revealed a similarity in the magnitude of difficulties and a decline in support directed toward mothers from primary care clinics due to a lack of incentives for staff and the cessation of structural support. Here, the “role of management and comprehensive institutional quality” in care and midwifery clinics within hospitals emerges as a key moderator and determinant of public health outcomes; contemporary literature in Sana’a on Total Quality Management (TQM) and the application of patient-centered standards (JCI) in Yemeni hospitals demonstrates that the implementation of quality standards, reliable systematic processes, clear policies, and midwife training has a direct and decisive impact on improving nursing staff performance and patient safety (Al-Saleet et al., 2025; Qabban & Al-Wesabi, 2025; Qabban et al., 2026; Alsalit et al., 2026). When medical facilities adhere to sound quality standards, midwives take the initiative to provide comprehensive and continuous nutritional counseling services and monitor each mother’s actual behavioral and dietary patterns, and ensure they receive regular, free supplements and iron, which contributes effectively, both administratively and professionally, to overcoming the logistical and material obstacles faced by mothers, improving their vital signs and clinical indicators, and preventing the deterioration of obstetric care (Shamlan & Al-Wesabi, 2026).

## 7. Conclusions, Recommendations, and Future Directions

### General Scientific Conclusions

This rigorous analytical field study of pregnant women in Sana’a for the year 2026 leads to the formulation of the following general conclusions:

- The pregnant mother’s nutritional awareness and practices contribute to explaining and determining approximately one-fifth (20.2%) of the total variance in maternal and fetal health and clinical outcomes and indicators in conflict settings.
- The mother’s actual physical implementation and behavioral practices have a stronger and more decisive physiological and obstetric predictive power in determining the clinical reality of the mother and fetus compared to merely possessing theoretical knowledge.
- Normal levels of fetal development, growth, and estimated weight mask severe biological depletion and exhaustion of the mother’s cellular and mineral stores, which clinically manifest as her weakness, anemia, compromised immunity, and deteriorating psychological and emotional state.
- Sociological and functional determinants (the mother’s high educational level, the family’s stable monthly income, and the occupation of homemaker) act as

protective shields and barriers that reinforce nutritional behavior and health outcomes, whereas poverty and ignorance represent structural obstacles that hinder the translation of awareness into practical behavior.

- The disappearance of statistical differences according to the stage of pregnancy and its current ranking scientifically confirms that the accumulation of a culture of healthy dietary behavior requires intentional education and organized clinical guidance through care channels; it is not generated automatically or innately through the repetition of previous births.

### Procedural and Practical Recommendations for Decision-Makers

Based on the results, conclusions, and formulation of the model’s causal relationships, this study presents the following procedural and practical recommendations to meet the requirements of international arbitration and inform decision-makers:

### First: To the Ministry of Public Health and Population and international organizations (UNICEF, WHO, UNFPA)

- Implementing the “Crisis-Sensitive Economic Nutrition Education” strategy through a radical shift in guidance campaigns—from formulating traditional guidelines based on expensive food groups (such as fresh meat, fish, and imported cheeses) to providing a national educational guide focused on “low-cost, nutritionally valuable local food alternatives” (such as combining legumes like fava beans and lentils with grains to form a complete protein, and consuming liver and dark green leafy vegetables for iron, and sesame and its sources for calcium), to overcome the barrier of declining purchasing power faced by 71% of pregnant women.
- Integrate psychological and physiological support into prenatal care by monitoring and managing symptoms of nausea, morning sickness, and psychological stress (through vitamin B6 supplements and psychological-nursing support) as critical preventive measures to address the most significant barriers to proper nutrition (80%).
- Targeting in-kind food basket assistance to the most vulnerable mothers: Based on observed disparities, in-kind food support and direct supplements should be allocated to illiterate mothers, the poorest and most financially disadvantaged mothers, and working mothers to reduce behavioral and health gaps among them.
- Developing crisis-sensitive nutrition education strategies centered on quality care and midwifery based on effective behavioral empowerment.

### Second: For midwives, nursing staff, and hospital administrations in Sana’a

- Commit to implementing patient-centered quality-of-care standards by training midwives in effective behavioral communication skills and assessing each mother’s actual dietary history rather than relying solely on a quick medical examination, while ensuring the distribution of free iron, folic acid, and iodine supplements.
- Launch and activate participatory support clubs and groups for pregnant women to organize cooking workshops and practical demonstrations on preparing affordable and accessible healthy meals, and to combat the use of qat, “bika,” and secondhand tobacco smoke.

### Proposals for Promising Future Research Directions

To address the limitations of the current study, researchers at Yemeni universities are encouraged to direct their research efforts toward the following promising applied topics:

· “A Longitudinal Cohort Study to Measure the Impact of Maternal Awareness Levels and Dietary Practices on Neonatal Neurodevelopmental Outcomes in Yemen”: A longitudinal study tracking mothers from the first trimester of pregnancy through six months postpartum to monitor the impact of maternal nutrition on infant cognitive indicators.

· “Causal Modeling and Mixed Qualitative-Quantitative Analysis of Pica and Its Relationship to Micronutrient Deficiency and Psychological Anxiety Among Pregnant Women in Sana’a”: An in-depth specialized study to assess blood levels of iron and zinc and their behavioral and biological relationship to the consumption of non-food items under conflict conditions.

· “The Impact of Implementing Total Quality Management (TQM) Standards in Maternal Care Clinics on Improving Nutritional Practices and Reproductive Health for Mothers in Government Hospitals in Sana’a”:

A. clinical-administrative study to measure the impact of improved midwife performance and health policies on the quality of nutritional counseling and the health of pregnant women.

## 8. Ethical and Administrative Considerations (Declarations)

### Ethical Considerations

The present study obtained ethical and professional approval from the Ethics Committee for Medical Research and the Deanship of Environment and Community Service on September 21, 2021, at the Faculty of Medical and Applied Sciences, Republic of Yemen. Strict adherence was maintained to obtaining voluntary informed consent (verbal and written) from all pregnant women participating in the field sample prior to completing or being interviewed for the questionnaire. The researchers ensured the absolute confidentiality of individual participants’ data and stored it in a secure environment designated solely for academic research purposes, while guaranteeing mothers complete freedom to withdraw from the study at any time without providing reasons.

## Data Availability

The raw data underlying this study contain sensitive clinical and socio-economic information of pregnant women living in a conflict setting. Due to ethical restrictions imposed by the Ethics Committee for Medical Research and the Deanship of Environment and Community Service at 21 September University for Medical and Applied Sciences, making the raw dataset publicly available is prohibited to protect participant confidentiality. However, the minimal data set can be made available to qualified researchers upon reasonable request to the Deanship of Graduate Studies and Scientific Research at 21 September University via the official institutional.

## Funding and Acknowledgments

This research was conducted and completed in the field and statistically analyzed entirely through self-funding.

The researchers extend their sincere thanks, appreciation, and deep gratitude to the Presidency of 21 September University for Medical and Applied Sciences, the Deanship of Environment and Community Service, and the Medical Training and Rehabilitation Center for their continuous support. We also extend our thanks to the midwifery students who participated in distributing and collecting the questionnaires from the participating mothers, and we express our gratitude to all the pregnant mothers who gave of their time and effort to participate and enrich the field questionnaire.

